# APOE E4 GENOTYPE PREDICTS SEVERE COVID-19 IN THE UK BIOBANK COMMUNITY COHORT

**DOI:** 10.1101/2020.05.07.20094409

**Authors:** Chia-Ling Kuo, Luke C Pilling, Janice L Atkins, Jane AH Masoli, João Delgado, George A Kuchel, David Melzer

## Abstract

The novel respiratory disease COVID-19 produces varying symptoms, with fever, cough, and shortness of breath being common. In older adults, we found that pre-existing dementia is a major risk factor (OR = 3.07, 95% CI: 1.71 to 5.50) for COVID-19 severity in the UK Biobank (UKB). In another UK study of 16,749 patients hospitalized for COVID-19, dementia was among the common comorbidities and was associated with higher mortality. Additionally, impaired consciousness, including delirium, is common in severe cases. The ApoE e4 genotype is associated with both dementia and delirium, with the e4e4 (homozygous) genotype associated with a 14-fold increase in risk of Alzheimer's disease compared to the common e3e3 genotype, in populations with European ancestries. We therefore aimed to test associations between ApoE e4 alleles and COVID-19 severity, using the UKB data.

---

The novel respiratory disease COVID-19 produces varying symptoms, with fever, cough, and shortness of breath being common. In older adults, we found that pre-existing dementia is a major risk factor (OR = 3.07, 95% CI: 1.71 to 5.50) for COVID-19 severity in the UK Biobank (UKB) [1]. In another UK study of 16,749 patients hospitalized for COVID-19 [2], dementia was among the common comorbidities and was associated with higher mortality. Additionally, impaired consciousness, including delirium, is common in severe cases [3]. The *ApoE* e4 genotype is associated with both dementia and delirium [4], with the e4e4 (homozygous) genotype associated with a 14-fold increase in risk of Alzheimer’s disease [5] compared to the common e3e3 genotype, in populations with European ancestries. We therefore aimed to test associations between *ApoE* e4 alleles and COVID-19 severity, using the UKB data.

UKB is a community cohort currently aged 48 to 86 [6]. COVID-19 laboratory test results for UKB participants in England are available from March 16 to April 26, 2020, the peak period of COVID-19 incidence in the current outbreak. During this period COVID-19 testing was largely restricted to hospital in-patients with clinical signs of infection, and therefore test positivity is a marker of severe COVID-19 infection [7].

We analyzed UKB data from genetically European ancestry participants [8] (n=451,367, 90% of sample) attending baseline assessment centers in England (n=398,073), excluding participants who died before the epidemic (n=15,885). Single nucleotide polymorphism (SNP) data for rs429358 and rs7412 was used to determine *ApoE* genotypes: *ApoE* e4e4 homozygotes (n= 9,022, 3%), e3e4 (n= 90,469, 28%), and e3e3 (most common genotype, n= 223,457, 69%) genotype groups (final n=322,948). Mean age was 68 years (SD= 8) with 176,951 females (55%). There were 622 positive COVID-19 patients (Table 1) including 37 with e4e4 (positivity rate: 410/100,000) and 401 with e3e3 (179 per 100,000). A logistic regression model was used to compare e3e4 or e4e4 genotypes to e3e3 for COVID-19 positivity status, adjusted for: sex; age at the COVID-19 test or age on 26th April, 2020 (the last test date); baseline UKB assessment center in England; genotyping array type; and the top five genetic principal components (accounting for possible population admixture).

**Table 1.** Risk of severe COVID-19, comparing participants with *ApoE* e3e4 or e4e4 to e3e3 genotypes in UK Biobank.

|  | n | Negative or<br>not tested | Positive | Positivity rate<br>per 100,000 | OR (95% CI)* | P-value |
| --- | --- | --- | --- | --- | --- | --- |
| All |  |  |  |  |  |  |
| e3e3 | 223,457 | 223,056 | 401 | 179 | - | - |
| e3e4 | 90,469 | 90,285 | 184 | 203 | 1.14 (0.95, 1.35) | 0.15 |
| e4e4 | 9,022 | 8,985 | 37 | 410 | 2.31 (1.65, 3.24) | 1.19E-06 |
| Excluding dementia |  |  |  |  |  |  |
| e3e3 | 222,968 | 222,574 | 394 | 177 | - | - |
| e3e4 | 90,013 | 89,840 | 173 | 192 | 1.09 (0.91, 1.31) | 0.338 |
| e4e4 | 8,877 | 8,840 | 37 | 417 | 2.39 (1.71, 3.35) | 4.26E-07 |
| Excluding hypertension |  |  |  |  |  |  |
| e3e3 | 151,018 | 150,792 | 226 | 150 | - | - |
| e3e4 | 61,249 | 61,157 | 92 | 150 | 1.00 (0.79, 1.28) | 0.981 |
| e4e4 | 6,120 | 6,098 | 22 | 359 | 2.41 (1.56, 3.74) | 8.21E-05 |
| Excluding coronary artery disease |  |  |  |  |  |  |
| e3e3 | 204,017 | 203,684 | 333 | 163 | - | - |
| e3e4 | 82,099 | 81,948 | 151 | 184 | 1.13 (0.93, 1.37) | 2.070E-01 |
| e4e4 | 8,164 | 8,132 | 32 | 392 | 2.43 (1.69, 3.50) | 1.65E-06 |
| Excluding Type II diabetes |  |  |  |  |  |  |
| e3e3 | 211,482 | 211,136 | 346 | 164 | - | - |
| e3e4 | 85,983 | 85,827 | 156 | 181 | 1.11 (0.92, 1.34) | 0.275 |
| e4e4 | 8,616 | 8,581 | 35 | 406 | 2.51 (1.77, 3.55) | 2.42E-07 |

\* adjusted for sex, age at the COVID-19 test or age on 26<sup>th</sup> April, 2020 (the last test date), assessment center in England, genotyping array type, and the top five genetic principal components

*ApoE* e4e4 homozygotes were more likely to be COVID-19 test positives (Odds Ratio = 2.31, 95% Confidence Interval: 1.65 to 3.24, p= 1.19×10^−6^) compared to e3e3 homozygotes (Table 1). The association was similar after removing participants with *ApoE* e4 associated diseases that were also linked to COVID-19 severity: participants without dementia (OR= 2.39, 95% CI: 1.71 to 3.35); hypertension (OR= 2.41, 95% CI: 1.56 to 3.74); coronary artery disease (myocardial infarction or angina) (OR= 2.43, 95% CI: 1.69 to 3.50) or type 2 diabetes (OR= 2.51, 95% CI: 1.77 to 3.55) (Table 1), based on pre-existing diagnoses from baseline self-reports or hospital discharge statistics (updated to March 2017). The estimates were little changed using 136,146 participants with additional general practice data (up to 2017): participants without dementia (OR= 2.53, 95% CI: 1.46 to 4.39); hypertension (OR= 2.67, 95% CI: 1.34 to 5.32); coronary artery disease (OR= 2.86, 95% CI: 1.65 to 4.98) or type 2 diabetes (OR= 2.73, 95% CI: 1.57 to 4.76). The results were also similar after excluding 51,430 participants related to the 3rd-degree or closer (OR= 2.34, 95% CI: 1.62 to 3.38). Of 622 included participants who tested positive for COVID-19, 417 (67%) were noted to the laboratory to be inpatients when the sample was taken: unfortunately, data on later admission to hospital is not available [7]. Including only those known to have been inpatients when tested made little difference to the excess risk associated with *ApoE* e4e4 status (OR=2.32, 95% CI: 1.54 to 3.29), compared to OR=2.31 (95% CI: 1.65 to 3.24) using all the tested samples.

In conclusion, the *ApoE* e4e4 allele increases risks of severe COVID-19 infection, independent of pre-existing dementia, cardiovascular disease, and type-2 diabetes. *ApoE* e4 not only affects lipoprotein function (and subsequent cardio-metabolic diseases) but also moderates macrophage pro-/anti-inflammatory phenotypes [9]. The novel coronavirus SARS-CoV-2 causing COVID-19 uses the ACE2 receptor for cell entry. ACE2 is highly expressed in type II alveolar cells in the lungs, where *ApoE* is one of the highly co-expressed genes [10]. Further investigation is needed to understand the biological mechanisms linking *ApoE* genotypes to COVID-19 severity.

## Data Availability

This research was conducted using the UK Biobank resource, under application 14631. We thank the UK Biobank participants and coordinators for the dataset.

## Notes

### Competing Interest Statement

The authors have declared no competing interest.

### Funding Statement

CLK, LCP, GAK, and DM are supported by a R21 grant (R21AG060018) funded by National Institute on Aging, NIH. UK Medical Research Council award MR/S009892/1 (PI Melzer) supports JLA. JAHM is supported by NIHR Doctoral Research Fellowship DRF-2014-07-177. The views expressed in this publication are those of the author(s) and not necessarily those of the NHS, the National Institute for Health Research or the Department of Health.

